# Primary care provider preferences on dementia training and care navigation services: A qualitative study

**DOI:** 10.1101/2022.10.11.22280973

**Authors:** Jaime Perales-Puchalt, Kelsey Strube, Ryan Townley, Michelle Niedens, Hector Arreaza, Jana Zaudke, Jeffrey M Burns

**Affiliations:** University of Kansas Alzheimer’s Disease Research Center, Fairway, KS; University of Kansas Medical Center, Kansas City, KS; University of Kansas Health System, Kansas City, KS; Clínica Sierra Vista, Bakersfield, CA; Rio Bravo Family Medicine Residency Program, Bakersfield, CA; Caritas Clinics, Kansas City, KS

**Keywords:** Education, dementia, healthcare professionals, Attitude of Health Personnel

## Abstract

**Background:** Dementia has no cure but interventions can stabilize the progression of cognitive, functional, and behavioral symptoms. Primary care providers (PCPs) are vital for the early detection, and long-term management of these diseases, given their gatekeeping role in the healthcare system. However, PCPs rarely implement evidence-based dementia care due to barriers such as limited dementia knowledge and time. Training PCPs and linking them to dementia care navigators may address these barriers.

**Objective:** We explored the preferences of PCPs about dementia care training programs and dementia care navigation services.

**Methods:** We conducted qualitative interviews with 24 PCPs recruited nationally via snowball sampling. We conducted all interviews via videocall and organized the transcripts for qualitative review to identify codes and themes, using a pragmatic approach, a qualitative description methodology, and thematic analysis methods.

**Results:** PCP preferences varied regarding the topic, duration, materials, modality, and incentives of the dementia training. With regards to dementia care navigation services, preferences varied with respect to whether they benefited the PCP or the patient, and which were the optimal qualities of a dementia care navigator.

**Conclusions:** Dementia training and care navigation services would benefit from embedding cultural proficiency within their content, materials, and navigation abilities. EMR-based decision-support tools would facilitate PCPs’ implementation of evidence-based dementia care.

## INTRODUCTION

Alzheimer’ s disease and related disorders (ADRD) pose a serious public health threat worldwide. ADRD is among the leading causes of mortality and disability among people in the US [1]. No treatment can currently prevent or stop the progression of ADRD. However, interventions can stabilize and delay the progression of cognitive, functional, and behavioral decline, improving outcomes for individuals with ADRD and their families [2, 3]. The healthcare system is well-suited to manage ADRD care, as most older adults in the US are insured and have a primary source of healthcare [4, 5]. Primary care providers (PCPs) are vital for the early detection and long-term management of these diseases given their gatekeeping role in the healthcare system [6].

Primary care and healthcare systems in general rarely implement evidence-based ADRD care. Healthcare providers rarely promote early detection via regular screening among those 65 and older [7]. In the setting of clear cognitive concerns, half of healthcare providers do a full workup to determine the cause [7, 8]. Even when abnormalities are found, half of individuals with ADRD are not given an official diagnosis by their provider [9]. Only half of individuals with ADRD receive medications for their disease [10]. Most family caregivers report unmet needs in at least one service area (e.g., activities of daily living, ADRD symptoms, time of the day in which care is required), and nearly one-third do not receive any type of caregiver support services [11, 12]. Barriers to the implementation of evidence-based ADRD care in healthcare include providers’ limited knowledge about its benefits, lack of necessary tools, and limited time to provide ADRD care [13, 14].

Training PCPs and linking them to ADRD care navigators can address some of the barriers to ADRD care implementation. Training can increase knowledge about the benefits of ADRD care, provide proper screening tools, and improve care management for PCPs. Care navigators are healthcare professionals or trained non-professionals that facilitate access to health and social services among patients and their families, facilitate continuity of care; and identify and remove barriers to care [15]. ADRD care navigators may assist with care coordination, making PCP visits more efficient. While some ADRD care training programs for PCPs and ADRD care navigation models exist [16-20], most are built from the top-down. Tailoring training programs and care navigation services to PCPs preferences may improve implementation and acceptability [21]. The objective of the current manuscript is to explore the preferences of PCPs about ADRD care training programs and ADRD care navigation services. To achieve this goal, we interviewed PCPs from diverse backgrounds across the US. Findings will inform the development and refinement of ADRD care training for PCPs and care navigation services.

## MATERIALS AND METHODS

This study used a qualitative design and collected data via semi-structured interviews. The aim of this study was to explore primary care provider preferences regarding ADRD training and care navigation services. This study was part of a larger project that aimed to identify what ADRD care services are offered among Latino families in primary care and how they are delivered across a variety of settings. PCP eligibility criteria included being a current or recently working medical doctor, a doctor in osteopathic medicine, a nurse practitioner, or physician assistant, in a primary care setting in the US.

Participants were initially recruited using snowball sampling by first contacting a network of PCPs from the faculty include in the projects’ research team and later asking participants for additional PCPs. The interviews took place between November 2020 and February 2021. All interviews were conducted via secure videoconference, except for one, that was conducted over the phone. The University of Kansas Medical Center Institutional Review Board approved this project. Before the interview, all participants completed an informed written consent online either via their computers, tablets, or phones.

The first author, JPP, interviewed all participants. The interview started with a short conversation aimed at developing rapport and explaining the main goals of the interview. Initial questions focused on characteristics of the participants and the clinics they worked in. Core questions of the survey asked about participants’ experience with the Latino families with ADRD they serve. The interviewer also asked participants to describe what an ideal ADRD training and integration of care navigation services would look like. The interviewer audiotaped all interviews, which were designed to last 45-60 minutes. Interviews were in English or Spanish. A professional team transcribed all interviews and the interviewer reviewed them for accuracy. The research team compensated participant’ s time with a $40 gift card.

We organized the transcripts for qualitative review, using a pragmatic approach, a qualitative description methodology, and thematic analysis methods [22-24]. We coded the content of the notes using a word processing program by identifying codes. These codes were later categorized into themes and subthemes using a spreadsheet. Two researchers conducted independent reviews of the codes and resolved coding disagreements through discussion and consensus. To bring rigor and validity to the research process, the interviewer used active listening techniques during the interview aimed at confirming the information shared by the participants. The interviewer also emphasized the fact that participants were the experts in their experiences to reduce power differentials. An additional researcher, KS, participated in the data coding process.

## RESULTS

### Characteristics of the sample

Table 1 shows the characteristics of the 23 PCPs. Half of PCPs were women (56.5%), US-born (52.2%), Latino (43.5%) of diverse origins, and non-Latino White (43.5%). Most PCPs were from urban settings (78.3%). While different US regions were represented, the Midwest was the most common one (65.2%). Half of PCPs reported being able to communicate with their patients in Spanish (52.2%). Most PCPs were medical doctors (65.2%) and nurse practitioners (30.4%). The types of clinics PCPs worked at and the distribution of Latino patients by PCP varied widely. Thirteen participants reported having received specific training or exposure to caring for older adults and more specifically people with ADRD. This previous training and exposure were completed via residency rotation, research projects, Ph.D. program or geriatrics specialization, and previous jobs at nursing homes, hospice or assisted living facilities, and home care.

**Table 1.** Characteristics of the sample (n=23)

|  | **Primary care providers** |  |
| --- | --- | --- |
|  | **%** | **n** |
| Women | 56.5 | 13 |
| Ethnic/racial group |  |  |
| Latino | 43.5 | 10 |
| Non-Latino White | 43.5 | 10 |
| Non-Latino Black | 8.7 | 2 |
| Non-Latino Asian | 4.3 | 1 |
| US-born | 52.2 | 2 |
| Urban setting | 78.3 | 18 |
| Interview in Spanish | 21.7 | 5 |
| Region |  |  |
| Midwest | 65.2 | 15 |
| Northeast | 8.7 | 2 |
| West | 17.3 | 4 |
| South | 0.0 | 0 |
| Puerto Rico | 8.7 | 2 |
| Can provide services in Spanish | 52.2 | 12 |
| Type of provider |  |  |
| Medical doctor | 65.2 | 15 |
| Nurse practitioner | 30.4 | 7 |
| Physician assistant | 4.3 | 1 |
| Type of clinic |  |  |
| Private | 47.8 | 11 |
| Safety net and federally qualified | 34.9 | 8 |
| Academic | 13.0 | 3 |
| PACE | 4.3 | 1 |
| Percent of Latino patients by provider |  |  |
| Less than 29% | 30.4 | 7 |
| 30-74% | 30.4 | 7 |
| 75%+ | 39.1 | 9 |
| Minutes of recording: Total; Min-Max | 1162 | 35-73 |

### Themes

Table 2 shows the themes, and subthemes identified in the qualitative analyses. Training preferences had five themes: the topics they wanted to receive training in, the preferred duration of the training, the materials they would prefer to receive in their training sessions, the modality in which they would like to attend the training sessions, and motivations or incentives to attend. The ADRD care navigation services topic had three themes: the services they would like the navigator to provide to them as PCPs, and to their patients with ADRD, and the optimal qualities these navigators should have.

**Table 2.** Themes, subthemes, and related codes.

| Topic | Theme | Subtheme |
| --- | --- | --- |
| Training |  |  |
|  | Topic |  |
|  |  | Basic aspects of ADRD |
|  |  | ADRD detection |
|  |  | ADRD care |
|  |  | Social aspects of ADRD care |
|  | Duration |  |
|  |  | Specific time of day |
|  |  | Flexibility of the duration |
|  |  | Length and purpose of that length |
|  | Materials |  |
|  |  | Materials for PCPs |
|  |  | Materials for families |
|  | Modality |  |
|  |  | Technology-based |
|  |  | In person |
|  | Incentives |  |
|  |  | Cost-benefit other than learning |
|  |  | Awareness of incentives |
| ADRD care navigation services |  |  |
|  | PCP services |  |
|  |  | General interest for navigator services |
|  |  | Linking to services |
|  |  | Available via EMR |
|  |  | Assisting with intermediate treatment steps |
|  | Patient services |  |
|  |  | Linking to services |
|  |  | Address needs in general |
|  |  | Address medical needs |
|  |  | Address social and contextual needs |
|  | Optimal qualities |  |
|  |  | Convey services menu clearly |
|  |  | Assigned navigator |
|  |  | Accessibility |
|  |  | Cultural competency |
|  |  | Sample models of navigation |

#### Topic 1. ADRD training

Theme 1. Training topic. PCPs mentioned several topics they would like the ideal training to include. These include basic aspects of ADRD such as the biological basis, the types, common needs, and the latest research on the topic. Other topics included ADRD detection such as the need to receive training on early detection, interpreting neuroimaging, and diagnosing ADRD. Further topics included, best care practices in pharmacological interventions, behavioral interventions, and disease management. A PCP summarized these subthemes as follows: *“What I would really value is the most up-to-date information in the world of diagnosis, treatment, and making sure that I am not falling behind from the cutting edge”*.

Another topic included social aspects of ADRD care such as receiving training in community living options, care transitions, financial assistance, driving and home safety, referrals Alzheimer’ s Disease Research Centers, pharmacies, caregiver respite options including daycare or in-home care, and support groups. This topic also included training in how to handle patient referrals with different insurance statuses and improving cultural competence. A non-Latino White PCP said: *“I think just in general cultural competence. There are so many cultures that I feel like I am doing my best, but I am not. The thing that I struggle with the most as a PCP is making sure that the care that I provide is culturally relevant. It is hard to me as a Caucasian provider… I know some things, but I think a lot of times we also make assumptions incorrectly. Understanding our own personal biases and how that can clog pictures and assumptions that we make. So, I think the more cultural training we can have the better”*.

Theme 2. Training duration. Participants often described their preferred training duration. PCPs suggested that the training session took place at specific times of the day when they had allotted time to take courses, or at lunchtime. PCPs mentioned the benefit of having flexible training modules to help fit training programs to different personal preferences and organizational cultures. For example, a PCP mentioned that having a wide array of duration and timing options would be useful. Some PCPs thought training several professionals at the clinic all at once rather than specific individuals could be a better approach. A PCP said:

*“I know all of the organizations do like where they have quarterly provider meeting with all other providers. This past year we have been doing Zoom, but previously we would all meet up at the main location from all the satellites and we reserve like 45 minutes for a lecture or a specialist to come in, or someone who we can partner with, for services or education. That would be really cool if something like that where they set it up with that organization to get the information out to a lot of providers all at once”*.

Regarding the training length, most participants emphasized their busy schedules and their need for the training to last one or fewer days, ranging from 30 minutes to 8 hours. Few PCPs mentioned a higher number of days, but warned about the potential burden on their work:

*“In terms of how many sessions, if you do too many, people are not going to do them, so you need to do two, three, four or maybe five at the most”*.

Some of the PCPs who preferred more than one day of training, did so because they wanted future sessions for feedback after they had processed the information from the first session, and put what was learned into practice. Another reason for wanting more than one day of training was to get quick updates about the lessons. A PCP’ s point summarized others’ ideas too:

*“It might be good to do something where you do a deep dive and then you come back and you kind of renew the touched points and*… *I could see three trainings. Maybe one longer and then other shorter ones*… *Maybe with some type of activity in between*… *something to get it in there because I do better if I am given information, I process it and practice it, and try it out and then I review it again in some way shape or form and practice it”*.

Theme 3. Training materials. Some PCPs referred to materials the training could hand out. Materials for PCPs include an evaluation to test the knowledge the PCPs gained during the training. Several PCPs would like organized lists of assessment tools, and resources to use in their practice. PCPs stressed the importance of having a list of partners for referrals and consulting (e.g., neurologists, neuropsychologists, psychiatrists, community organizations), especially given the long waiting times in some specialty clinics. A popular need among PCPs was a decision support tool that would specify the steps PCPs need to follow to provide comprehensive ADRD care. This decision support tool may include condensed and expanded information options to reduce PCP burden and could be either integrated physically in their offices or into their EMRs. However, PCPs warn against receiving too many materials:

*“If we just are hanging it in places our goal to work gets so obnoxious, I walk by and then I get anxiety just there is so much stuff hanging there”*.

PCPs wanted to receive materials for their patients with ADRD and their caregivers during their training. The content of such materials could include what ADRD is, what to expect, how to provide care for a loved one, how to improve home safety, and discussions about transfers to different levels of care. Materials need to be in the appropriate language and literacy level. This last point is important, as even though PCPs tend to have some information to hand out in Spanish, there is doubt in their usefulness given the complexity of the information:

*“We struggle on the patient handout information component. If those things were made available and were made available in a bilingual format it would be very helpful”*.

*“I think patients like to have something they can hold on to that is understandable and easy… sixth grade reading level”*.

Theme 4. Training modality. Participants discussed their preferred modality for training sessions. PCPs suggested technology options such as integrating videos and open forums in their training, where trainees can follow up with further questions after the training. Most PCPs evoked technology-based training modalities that were remote or online. In fact, given potential time incompatibilities, some PCPs argued for pre-recorded sessions to help manage their times better. Most PCPs liked both remote and in-person formats. However, four PCPs preferred in-person training sessions, and cautioned against virtual and pre-recorded training formats.

*“I have a love/hate relationship with Zoom. I love that sometimes I can be able to do recorded lectures on my own time. The problem is, if I do not go out of my office… everybody knows I am here, and they bug me. So, even though I have scheduled time to attend these virtual conferences, they don’ t work, even if I am working from home, I just get bombarded. Whereas when I had to travel to something before, people tended to leave me alone because they know that I am there. Finding time to do them virtually, there is always some other thing that comes up that just demands my attention quicker. I like to do things when people ask me questions and really engage. With prerecorded lectures, there is a good chance I will not pay attention, I am just being honest here. I will engage for a little while, but then it is always going to draw me away, so the more it is interactive, the more it is engaging. I think that really helps”*.

Theme 5. Training incentives. Some PCPs talked about reasons, motivations, and incentives to attend training sessions other than learning. Several PCPs thought offering continuing education credits would make the training program more appealing. When asked about the possibility of offering training for free, a PCP mentioned that cost is usually not an issue, because they are assigned a yearly stipend for continuing education training. Some PCPs highlighted the importance of promotion to make PCPs aware of the training programs. For example, a PCP that practices in a hospital that has been offering a training program for two years said:

*“No, I have not attended the training yet. It is sad to say that, but as a PCP within the academic hospital, I am really just starting to become more and more aware what is available as far as resources, and that is sad to say because we are not a huge hospital, so I think definitely promoting that more would be really helpful”*.

#### Topic 2. ADRD care navigation services

Theme 1. PCP services. PCPs mentioned services that would benefit their practice. Several PCPs manifested an interest in having a care navigator available to them that specialized in ADRD. PCPs highlighted the importance of potential care navigators’ role in linking PCPs to services, by creating a community network of social services and clinical partners. The ADRD navigator would provide PCPs with lists of resources tailored to the needs of the patient and would help improve the communication between specialists and PCPs. Regarding this last point, a PCP said:

*“A ADRD care navigator would be helpful, because if I send the patient to a consultation with a neurologist, I would like to have the reports of the specialists and the laboratory tests they have done. I am interested in the specialists’ opinion because we do have that barrier as well, that we do not have the same EMR systems, so receiving the information from them for me sometimes is hard”*.

Some PCPs thought ADRD care navigators should be available via EMRs to make communication with them easier. Several PCPs talked about how care navigators could help by assisting with intermediate treatment steps. Help with intermediate steps would take the burden off them and allow them to focus on more aspects of care. Intermediate steps include completing intake forms, medicine lists, addressing social determinants of health, or getting the information PCPs need to follow up with their patients. A PCP described the potential for ADRD care navigators to reduce their burden as follows:

*“I think the big thing with primary care is just understanding that it is a flood and anything that can reduce the flood for us and make it really just hitting the high points, that would be incredibly helpful”*

Theme 2. Patient services. A common theme was the services PCPs thought ADRD care navigators could offer to their patients. Most PCPs saw great potential for care navigators in linking patients and their families to services for their clinical and social needs in a centralized way. Given cognitive issues and potential family burden, several PCPs stressed the need to have the care navigator assist in calling and communicating with those services, coordinating visit schedules, and reminding them of their appointments, in their preferred language. A PCP said:

*“Care navigators would be helpful by becoming the coordinating care point. That is some of the biggest things, just making sure that people get back with their appointments and following up with us. A lot of patients with memory issues forget. A lot of family members are working full-time as well. Being able to coordinate with their schedule so that they can bring their family member, making sure that they are taking advantage of the community resources”*.

PCPs mentioned different kinds of patient needs that care navigators could address. First, several PCPs stressed the need to assess unmet needs and address them in general. This was especially important given that PCPs did not feel they could address all the needs by themselves. A care navigator with a medical background could help assist with medication assistance. This includes keeping medication lists updated, medication reminders, contacting their healthcare provider or pharmacy if they need any medication. Additional roles included arranging transportation and logistics for clinic visits and reminding families of their clinic appointments. PCPs also thought care navigators could help address mood management by checking in on patients’ mood, providing emotional support, and having structured plans for when the patient is struggling. Regarding social and contextual needs care navigators could help with care transitions, risk prevention and safety, social determinants of health, and supporting the family. A comment that summarized some of these social and contextual needs is the following:

*“It would be good if navigators could be able to deal with family members. Family members have their own things going on too. It might be helpful having that person… may be a good liaison for caregivers too for social and emotional support, connecting them with therapists or with their own PCP or whatever they might need, also coordinating with like social work making sure people are not out of food, transportation…”*

Theme 3. Optimal qualities. PCPs mentioned optimal qualities that the ADRD care navigators should have. According to PCPs, an important care navigator quality would be explaining clearly and ahead of time what services they can provide, as well as the purpose and expected benefits. PCPs valued the idea of one specific navigator being assigned to the PCP, to promote consistency and trust. Several PCPs also highlighted the need for care navigators to be accessible when needed and via different formats (home visits, phone calls, clinic, virtually). PCPs emphasized the need for care navigators to be culturally and linguistically competent. This was especially necessary among Latinos, given the high percentage of patients and families who only speak or prefer Spanish. Three PCPs referred to existing models of care navigation in local cancer centers as they described their preferred qualities.

*“At the cancer center there is a navigation mechanism, usually a person, which is wonderful. I’ m able to call there and actually talk to a human being who will talk to my patient who just got the diagnosis of breast cancer and navigate them. That is a huge thing because then I know I’ m handing off my patient, I actually know who’ s going to call them. Navigation for me is key because it’ s such a complex experience trying to move through this broken system”*.

*“For the oncology department, they have nurse navigators. They are super helpful. I would just talk to one of them and tell them what I need, and they are really available on the phone or via email or whatever, and they respond very promptly. And then again helping me answer any of these questions. So ‘ I’ m seeing somebody, this is what I think they need, so what help can you provide me?’ ”*.

## DISCUSSION

This study aimed to explore the preferences of PCPs about ADRD care training programs and ADRD care navigation services. To achieve this goal, we interviewed 23 primary care providers across the US. These participants were diverse with respect to their professional background, region, and primary language. Important findings include the preference to receive content about early detection, cultural competence, and social determinants. ADRD training would benefit from being well-promoted, flexible with regards to technology and durations, and include brief follow-up refreshers or practice feedback. Preferred materials include lists of clinical and social services partners, as well as EMR-based decision support tools, and culturally-tailored educational readings. Preferred features of ADRD care navigators include reducing PCP burden, addressing patients’ multiple needs, being an accessible point of contact for PCPs, and linguistically and culturally competent.

PCPs included preferences on training content and materials to promote early detection and building cultural competence. These topics are rarely included in ADRD care training [16]. There is evidence that early detection can bring about health and economic benefits to families with ADRD [25-29]. Early detection gives individuals with ADRD the opportunity to participate in their care planning and to receive efficacious interventions earlier [30]. Addressing ADRD disparities among ethnically and racially minoritized populations is a key component of the US ADRD plan [31]. According to the National Academy of Medicine, an implementable and efficacious way to reduce disparities is by training healthcare providers in cultural competence [32]. Using materials and referring to resources that are linguistically and culturally tailored may reduce the gap in ADRD detection and care among minoritized populations [33].

PCPs’ preference for relatively brief and flexible training programs that capitalize on technology is not surprising given their busy schedules [13, 14]. These preferences, however, contrast with the rather time-intensive, in-person, and rigid schedules of most existing training programs [16]. The opportunity to receive feedback from the instructors after PCPs put their learning into practice is reasonable given the importance of performance experience in skills learning [34]. The preference for promotion of the training and continuing education credits allow awareness and provide an additional extrinsic value to enrolling the training, both important for behavior change [35].

Research has shown that ADRD care navigation services can improve patient and caregiver clinical outcomes of depression, neuropsychiatric symptoms, quality of life, and institutionalization rates [18-20]. Some of these models used nurse practitioners [19], social workers [20], or trained unlicensed professionals [18]. In line with our findings, all these models assess the needs of families with ADRD and address them with education and support. Also, consistent with our PCPs’ preferences, the ADRD care navigators from existing models refer families with ADRD to community services and communicate with the PCP and other healthcare providers to coordinate care. PCPs suggested integrating ADRD care navigators into the electronic medical records. To our knowledge, only one of the three care models we have cited has integrated them, and one plans to do so [18-20]. Such integration may help facilitate care coordination between the care navigator and other members of the care team. PCPs wanted care navigators to clearly explain their functions. PCPs also wanted care navigators to perform intermediate tasks to reduce their workload. Performing these intermediate tasks has the potential to reduce PCPs’ time burden, and help address specific ADRD needs that PCPs might not have enough experience with [13, 14].

This study has limitations. While the ability to do remote interviews via videocalls increased our reach, it also led to some communication issues that affected the quality of the information we gathered. The sample size was relatively small and not probabilistic, and Physician Assistants and Doctors in Osteopathy were poorly represented. We attempted to include PCPs from as many settings as possible, but we were unable to partner with the Veterans Affairs due to low response rates and complicated bureaucracy. Altogether, these issues reduce the generalizability of this study’ s findings. As with most studies, participants had a willingness to participate, which biased the selection of the sample. We do not know how much their opinions and preferences compare to PCPs who decided not to participate.

This study has implications for future practice. Findings can inform the development and refinement of ADRD care trainings for PCPs and care navigation services. ADRD education providers should consider the preferences evoked by PCPs. Training formats that allow for future feedback after practicing what was learned during the first session may improve adherence to guidelines. EMRs may allow better integration of decision support tools compared to paper materials. Clinics may benefit from already existing culturally-tailored and free materials for ADRD caregiver support [36]. The creation of partnerships such as a Provider-Based Research Network might address the needs of many PCPs who need ADRD training and services. Our findings help inform the roles and tasks of care navigators, as they include the qualities PCPs value most. Future research should integrate and test these preferences into new training formats and compare them with existing training curriculums in cluster trials. Before testing their effectiveness, researchers might want to conduct a survey to quantify these preferences in a sample that is representative of PCPs in diverse settings.

## CONCLUSION

This study has identified PCP preferences with respect to ADRD training and care navigator characteristics. Some of them are already being implemented in existing ADRD training courses and navigator models. However, other preferences (e.g., cultural proficiency for training content, materials and care navigator abilities, EMR-based decision-support tools) are not common practice. The bottom-down process of integrating these preferences into future training courses might improve their effectiveness and ultimately improve the lives of many families with ADRD.

## Data Availability

All data produced in the present study are available upon reasonable request to the authors

## Acknowledgements

This work was supported by the NIH under Grants K01 MD014177, R24 AG063724, and P30 AG072973. We also thank those who participated in the interviews, those who promoted it, and everybody who has contributed directly and indirectly to this research.

## Statement for ethical approval

The University of Kansas Medical Center Institutional Review Board approved this project (STUDY00145615).

